# Impact of *APOE*, *Klotho* and sex on cognitive decline with aging

**DOI:** 10.1101/2024.07.20.24310745

**Authors:** Kengo Shibata, Cheng Chen, Xin You Tai, Sanjay G Manohar, Masud Husain

**Affiliations:** Nuffield Department of Clinical Neurosciences, University of Oxford, Oxford, UK; Division of Clinical Neurology, John Radcliffe Hospital, Oxford University Hospitals Trust, Oxford, United Kingdom; Department of Experimental Psychology, University of Oxford, UK

**Keywords:** aging, Alzheimer’s Disease, antagonistic pleiotropy, apolipoprotein ε, cognition, klotho, neurodegeneration

## Abstract

The effects of *APOE* and *Klotho* genes, both implicated in aging, on human cognition as a function of sex and age are yet to be definitively established. Here we showed in the largest cohort studied to date (*N =* 320,861) that *APOE* homozygous ε4 carriers had a greater decline in cognition with aging compared to ε3 carriers (ε4/ε3 & ε3/ε3) as well as smaller hippocampi and amygdala (*N =* 37,976). Critically, sex and age differentially affected the decline in cognition. Younger (40 - 50 years) female homozygous ε4 carriers showed a cognitive advantage over female ε3 carriers, but this advantage was not present in males. By contrast, *Klotho-VS* heterozygosity did not affect cognition or brain volume, regardless of *APOE* genotype, sex or age. These cognitive trajectories with aging demonstrate clear sex-dependent antagonistic pleiotropy effects of *APOE* ε4, but no effects of *Klotho* genotype on cognition and brain volume.

## Introduction

Several lines of evidence have suggested that genotype can influence the risk of cognitive decline and ultimately dementia^1,2^. However, the relative contribution of genetic and environmental risk factors on brain health continues to be vigorously debated^3,4^. Of all genes studied to date, the ε4 variant of the apolipoprotein E (APOE) gene has been most strongly identified as a significant risk factor for developing sporadic Alzheimer’s disease (AD)^1,5–7^. Even healthy carriers of the *APOE* ε4 gene have been reported to have accelerated cognitive decline compared to non-carriers^8–10^, with significant structural and functional brain alterations^11–13^ and localised effects on the hippocampus^14–17^. As these effects are observed in asymptomatic individuals, before the onset of either mild cognitive impairment (MCI) or AD, genotyping provides a potentially powerful means of stratifying which individuals are more likely to develop dementia^18^.

However, the literature on how *APOE ε4* influences the progression and manifestation of cognitive decline over the life span in healthy individuals remains mixed. Crucially, the directionality of the effects caused by carrying *APOE* ε4 on the brain has been perplexing, especially in younger individuals. Several studies have reported a cognitive advantage of carrying *APOE* ɛ4 at earlier ages^19–22^. These findings suggest that the *APOE* ε4 variant may confer an advantage in early adulthood, but this comes at the cost of a higher rate of cognitive decline and increased risk of dementia in later life. This pattern aligns with the concept of ***antagonistic pleiotropy***, where a gene that provides benefits in early life may lead to disadvantages with aging^23,24^. This early life benefit of *APOE* ε4 might account for its evolutionary persistence.

In addition to the significant impact of the *APOE* genotype on mediating cognitive decline, studies have also emphasised the potentially crucial role of an individual’s sex^25^. Females compared to male carriers of the ε4 allele are reported to be at a higher risk of developing AD^26–28^ and experience faster rates of memory decline in MCI^29^. A memory advantage in male *APOE* ε4 carriers in midlife (40 - 51 years old)^30^ and a cognitive disadvantage in female *APOE* ε4 carriers (50 - 80 years old)^31^ has also been reported. Therefore, the strong interaction between sex and age in cognitive resilience and decline underscores the importance of examining how sex differentially influences the impact of the APOE ε4 allele. However, it remains to be definitively established how the *APOE* genotype and age, in relation to an individual’s sex, affect cognition^32^.

In addition to *APOE*, another significant genetic factor considered important in influencing cognitive function is the *Klotho* genotype. The Klotho protein has been linked to increased lifespan when overexpressed in mice^33^, while mice lacking this protein lived for shorter periods^34^. Building on these findings, human studies have explored *Klotho*’s potential impact on brain health. Carrying one, but not two copies of the KL-VS haplotype, i.e., *Klotho-VS* heterozygosity (*Klotho* VS/FC) has been identified as a potential genetic factor with a protective effect, increasing longevity^35–37^ and crucially also improving cognition^38,39^. *Klotho* VS/FC has also been associated with larger volumes of the right dorsolateral prefrontal cortex, and corresponding enhanced executive function, not seen in *Klotho* homozygotes^40^.

Mechanistically, *Klotho* VS/FC has been associated with an increase in serum Klotho protein levels^41^ which has recently been shown to correlate with cognitive scores^42,43^. Critically, Klotho*- VS* heterozygosity has been reported to be protective against amyloid deposition caused by *APOE* ε4^44,45^. These *Klotho*-mediated genetic effects have motivated the investigation into treatments targeting the *Klotho* pathway. However, the effects of *Klotho* VS/FC are still under considerable debate^46–48^. Conflicting findings have been reported with one study reporting that *Klotho* VS/FC does not confer a cognitive advantage^49^ and another showing shorter, not longer, survival in heterozygous carriers^48^. Any benefits conferred by *Klotho genotype* may potentially be age or sex-dependent and therefore warrant further investigation in larger sample sizes.

Here, we sought to address the polygenic effects of *APOE* and *Klotho* on cognition and brain structures throughout aging using an extensive dataset from the UK Biobank, the largest sample size studied reported to date. Studying individuals before disease onset allows the assessment of risk factors in a healthy population to capture early signs of cognitive changes in the healthy state. We established a composite behavioural score derived from a battery of cognitive tasks. In addition, structural MRI was used to extract grey matter volumes of predefined brain regions as a metric for brain integrity. We aimed to separately assess the role of each gene variant across age and sex on a composite score of cognition and grey matter volumes. The large sample size afforded by the UK Biobank allowed us to establish 1) the ages at which any effects of *APOE* and *Klotho* on cognition or brain structure appear 2) whether any of these effects are sex-dependent and 3) the interaction between these two different genes. We hypothesised that *APOE* homozygous ε4 carriers would exhibit more rapid cognitive decline and neuroanatomical differences with aging, while individuals with *Klotho*-VS heterozygosity would exhibit protective effects against such decline^44,45^. By investigating these effects, we aimed to contribute to the current understanding of genetic predispositions to neurodegeneration in healthy individuals across sex and aging.

## Results

### APOE’s effect on cognition is age and sex dependent

To study the effect of *APOE* and *Klotho* genotype on the cognitive functions of healthy adults, a composite score was derived from nine cognitive tasks of 312,524 participants aged 40-70, by taking the first principal component (*See methods &* ***Table 2***). How this composite score of cognition varied with *APOE* genotype (ε33/34, ε44, ε22/23), *Klotho* genotype (FC/FC, VS/FC), sex (female, male), and age was assessed using multiple linear regression. This gave a factorial *3 x 2 x 2 x age* design, with effects visualised in **Figure 1 & 2**. Each main effect and interaction term without the age predictor reflected an effect at the baseline age of 40.

**Figure 1:**
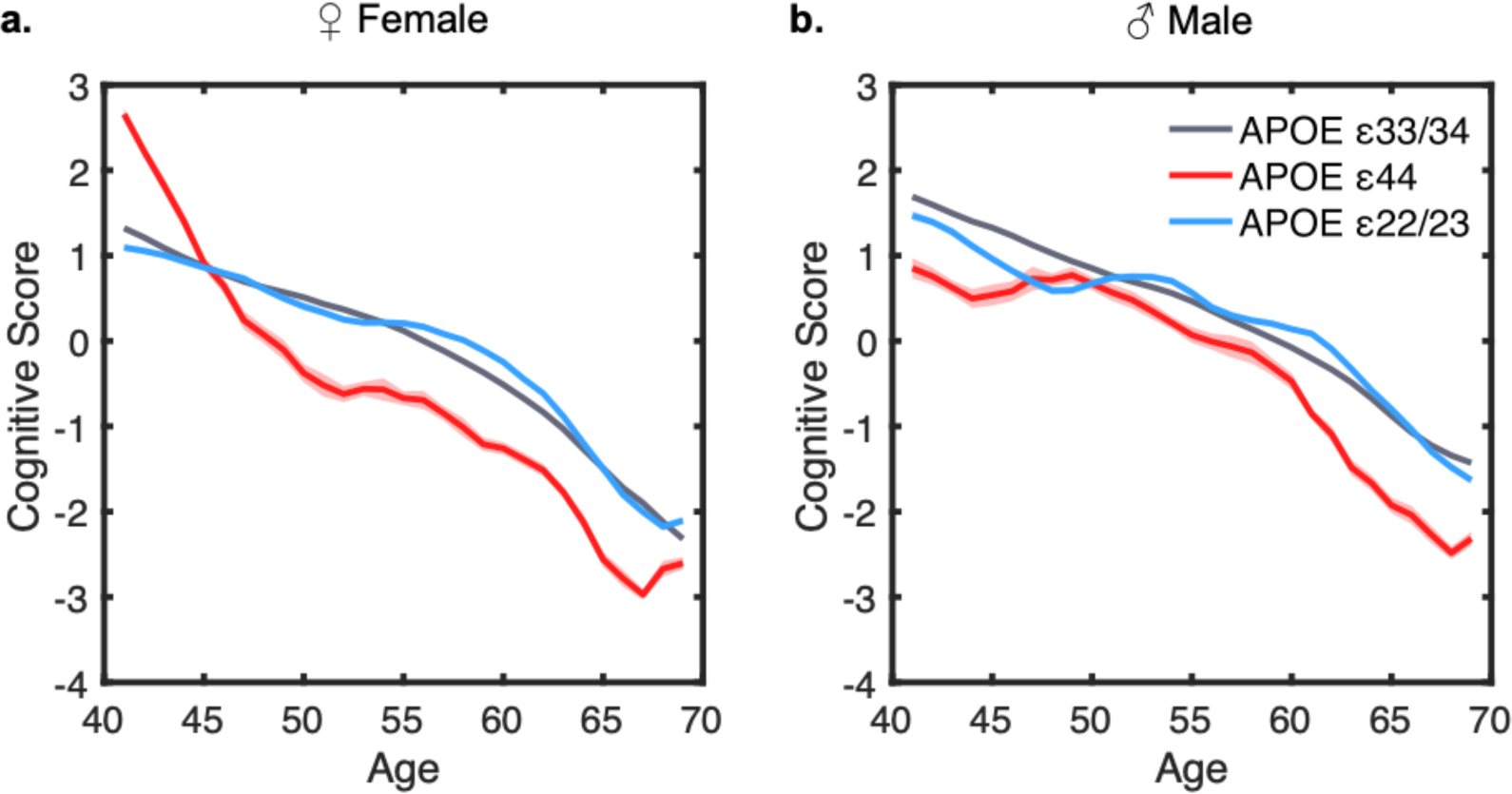
The influence of *APOE*, sex and age on cognitive performance. **Panel a (Female):** Comparison of cognitive composite scores among APOE ε33/34 (grey), ε44 (red), and ε22/23 (blue) genotypes revealed genotype and age-dependent effects on cognition. Female ε44 carriers demonstrated a cognitive advantage over ε33/34 carriers until age 45, followed by a steeper decline. In ε22/23 carriers, compared to ε33/34, a modest cognitive improvement was observed between the ages of 55 and 65. **Panel b (Male):** Cognitive composite scores for males indicate that *APOE* ε33/34 consistently outperformed ε44 across all age groups. ε22/23 carriers exhibit a slight cognitive improvement during middle age. Cognitive scores are represented using a three-year moving average with individual’s ages rounded to the nearest integer. Shaded area represents standard error of the mean (SEM) for each age group.

**Figure 2:**
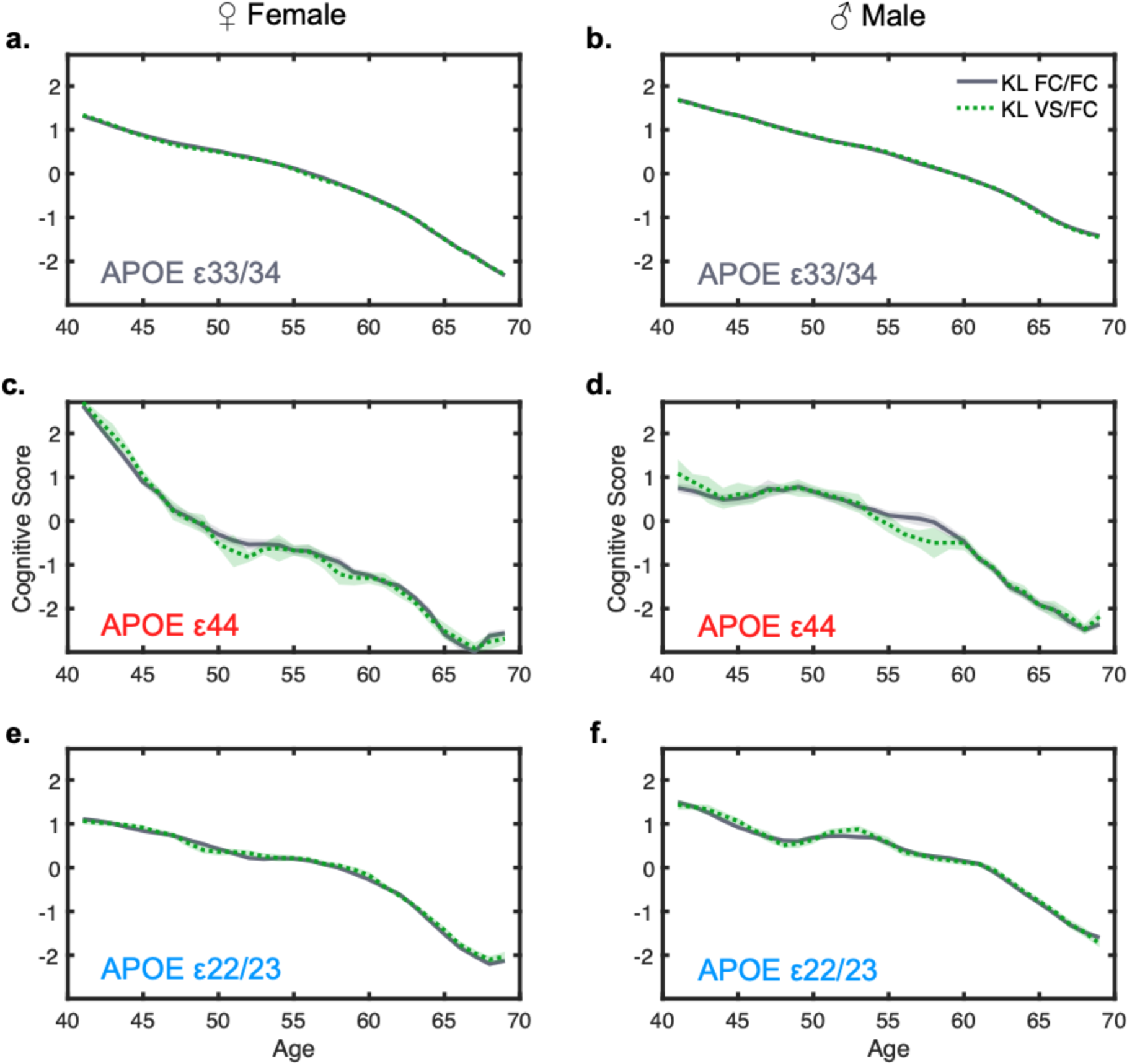
The influence of *Klotho*, sex and age on cognitive performance. **Panel a-f** (Left Panels Female, Right Panels Male): Comparison of cognitive scores between Klotho genotypes (Klotho FC/FC vs. FC/VS) showed minimal differences in cognition, regardless of APOE genotype. Contrary to expectations, the anticipated cognitive protection in APOE ε44 carriers by the Klotho VS/FC genotype was not evident, as shown in panels c & d. Cognitive scores are represented using a three-year moving average with individual’s ages rounded to the nearest integer. Shaded area represents the SEM for each age group.

At baseline age, carrying *APOE* ε44 positively affected cognition (β = 0.23, SE = 0.044, t = 5.21, p < 0.001) compared to ε33/34 (**Figure 1, panel a & b**). By contrast, carrying *APOE* ε22/23 negatively affected cognition (β = -0.094, SE = 0.020, t = -4.65, p < 0.001). No significant main effect of *Klotho-VS/FC* was observed (KL: β = -0.022, SE = 0.014, t = -1.58, p = 0.11, **Figure 2, panel a-f)**. Furthermore, *Klotho* status did not interact with sex (KL X sex: β = 0.030, SE = 0.021, t = 1.44, p = 0.15) nor *APOE* genotype (KL X *APOE* ε44: *Klotho* β = 0.052, SE = 0.081, t = 0.64, p = 0.52; KL X *APOE* ε22/23: β = -0.019, SE = 0.038, t = -0.49, p = 0.63) nor age (KL X age: β = 0.001, SE = 0.001, t = 0.79, p = 0.43). Therefore, the anticipated protective effects of *Klotho* VS/FC for APOE ε44 were not observed. The regression coefficients are plotted in **Figure 3**.

**Figure 3:**
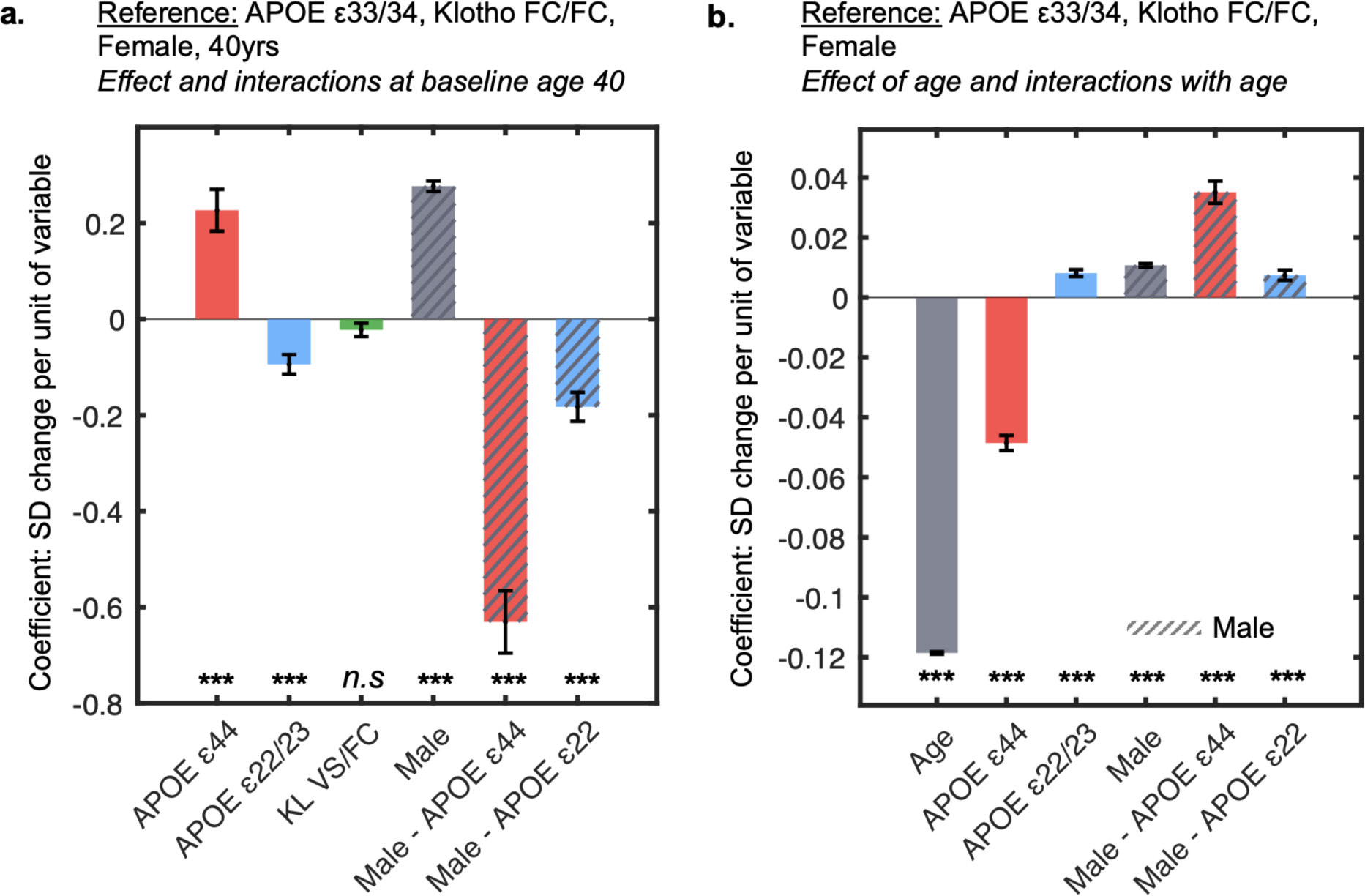
Multiple regression coefficients assessing the influence of *APOE*, *Klotho*, sex and age on cognition. **Panel a:** Coefficients from the multiple regression, representing the main effects and the interaction between sex and APOE genotype. Effects of sex and genotype on cognitive performance are shown, where reference variables are *APOE* ε33/34, *Klotho* FC/FC and female at baseline age of 40. **Panel b:** Coefficients from the multiple regression representing the effect of age on cognition, along with age interactions. Only statistically significant interactions are visualised. The cognitive composite score (units in SD) used is derived from the first principal component of cognitive tests from the UK Biobank. Error bars represent the SEM. Significance levels are denoted as follows: *p < 0.05, **p < 0.01, ***p < 0.001.

Across all genotypes, at the baseline age of 40, males showed significantly better cognition than females (sex: β = 0.28, SE = 0.011, t = 25.34, p < 0.001). However, the interaction between *APOE* and sex revealed that at age 40, males had substantially lower cognitive performance in ε44 (ε44 x sex: β = -0.63, SE = 0.065, t = -9.71, p < 0.001) and ε22 (ε22 x sex: β = -0.18, SE = 0.030, t = -6.04, p < 0.001) compared to females of the same genotype. To reiterate, the directionality reported here only considers effects at the baseline age 40 (**Figure 3, panel a**).

Age plays a crucial role in cognitive decline, as evident in the trajectories of cognitive performance in all panels (**Figure 1 & 2**). Accordingly, age strongly reduced cognitive performance (main effect of age: β = -0.12, SE = 0.00, t = -279.55, p < 0.001). The following analysis reports the predictors and their relationship with age (**Figure 3: panel b**). The cognitive *advantage* associated with the *APOE* ε44 genotype that was seen at baseline age was reversed in older participants. Indeed, the expected negative influence of *APOE* ε44 on cognition increased with age (age x ε44: β = -0.049, SE = 0.003, t = -19.07, p < 0.001), underscoring an age-dependent and accelerated cognitive decline in *APOE* ε44 carriers. For carriers of *APOE* ε22/23, the slope of the cognitive change across age was less steep (slower) with aging compared to ε33/34 carriers (age x ε22: β = 0.008, SE = 0.001, t = 7.00, p < 0.001).

A significant age-by-sex interaction revealed that females had a faster decline in cognition with aging overall (age x sex: β = 0.011, SE = 0.001, t = 17.23, p < 0.001). Critically, this decline with age in *APOE* ε44 carriers was depended on sex, with females being more strongly impacted by the gene (ε44 x sex x age: β = 0.035, SE = 0.004, t = 9.46, p < 0.001) (**Visualised in Supplemental Info 1**). The *APOE* ε22/23 effect with age, albeit weaker, was also modulated by sex, whereby the early female advantage declined faster with age (age x ε22/23: β = -0.18, SE = 0.030, t = -6.04, p < 0.001). The coefficients from the statistics are visualised in **Figure 3 panels a-b** and summarised in **Table 2**.

### Age determines the influence of *APOE* genotype

To evaluate the interactions between *APOE* genotype and sex in different age categories, we partitioned the analysis into three age categories, 40 – 50, 50 – 60 and 60 – 70 years. This segmentation was chosen to divide the age ranges into equal intervals to assess the age-related directionality of effects conferred by genes and sex on cognition. We excluded the *Klotho* variable from this follow-up analysis as it did not significantly predict cognition in the previously reported models. The positive effect of *APOE* ε44 on cognition in the 40-50 age group (ε44: β = 0.12, SE = 0.030, t = 3.78, p < 0.001), reversed to a negative effect in the subsequent age brackets; 50-60 (ε44: β = -0.85, SE = 0.030, t = -28.46, p < 0.001) and 60-70 years (ε44: β = -0.76, SE = 0.033, t = -22.61, p < 0.001) as illustrated in **Figures 4, panel a-c**. A similar pattern was observed for *APOE* ε22/23, showing a negative effect in the 40-50 age range (ε22/23: β = -0.073, SE = 0.014, t = -5.14, p < 0.001), which transitions to positive effects in the 50-60 (ε22/23: β = 0.100, SE = 0.014, t = 7.35, p < 0.001) and 60-70 (ε22/23: β = 0.091, SE = 0.015, t = 6.03, p < 0.001) age groups. The effect of being homozygous for *APOE* ε4 is positive at younger ages and is only detrimental above 50 years of age.

**Figure 4:**
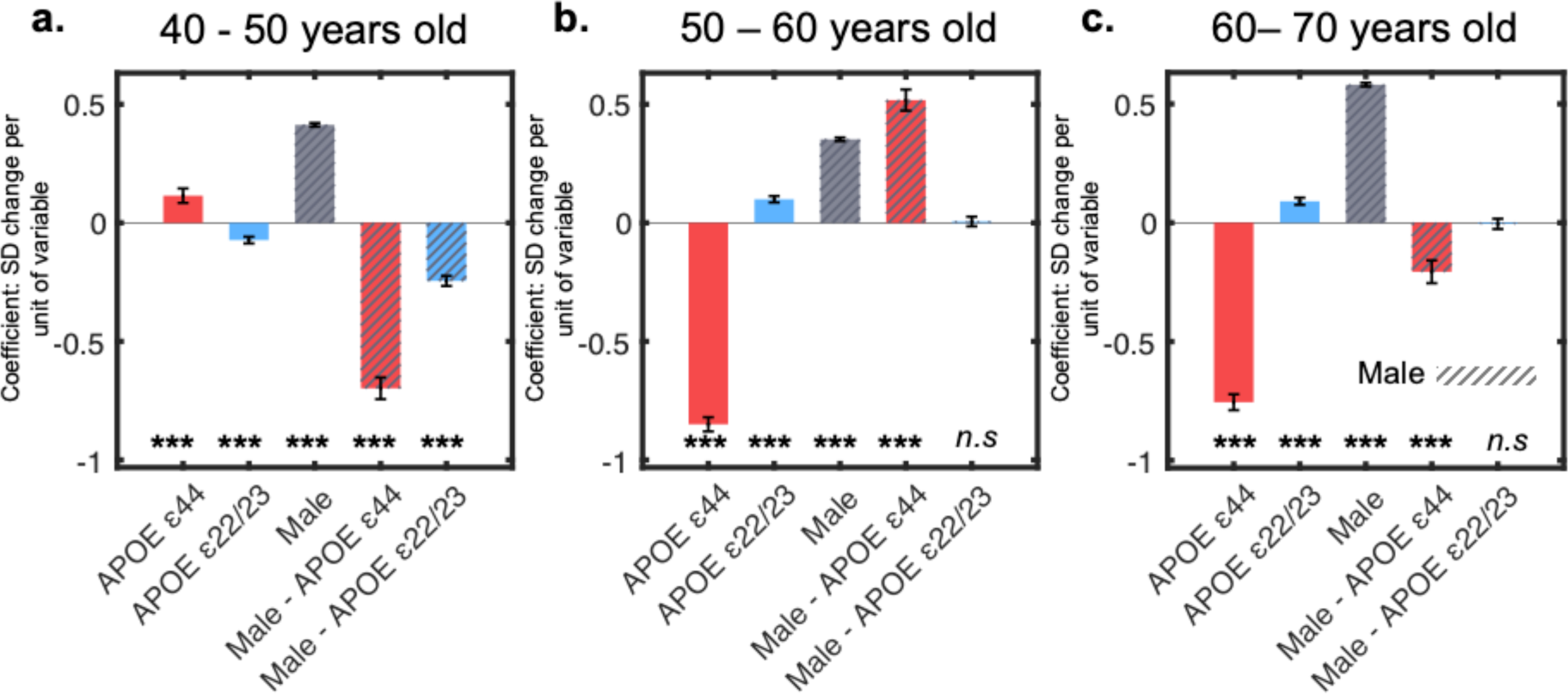
Effect of *APOE* and sex by age ranges. **Panel a-c:** Coefficient values for the main effects of *APOE* and sex, and their interaction, analysed through sub-sampling across three age groups: 40-50, 50-60, and 60-70 years. The *Klotho* genotype was not significant in the previous regression and thus excluded from these analyses. Error bars represent the SEM. Significance levels are indicated as *p < 0.05, **p < 0.01, ***p < 0.001.

Despite males generally displaying better cognition scores across all ages, a pronounced female advantage at ages 40-50 was shown by a significant interaction between sex and *APOE* ε44 (sex X ε44: β = -0.70, SE = 0.046, t = -15.27, p < 0.001) and *APOE* ε22/23 (sex X ε22/23: β = 0.52, SE = 0.045, t = 11.54, p < 0.001). However, this advantage was not apparent in the 50-60 age group (Male-APOE ε44 bar in dashed red). It re-emerged in the 60–70-year-olds with *APOE* ε44 females outperforming males (sex X ε44: β = -0.21, SE = 0.048, t = -4.32, p < 0.001). Although female APOE ε22/23 carriers had a cognitive advantage between 40-50 (sex X ε22/23: β = -0.25, SE = 0.021, t = -11.47, p < 0.001), sex no longer affected this effect at older ages (sex X ε22/23 - 50-60: β = 0.006, SE = 0.020, t = 0.28, p = 0.78; sex X ε22/23 - 60-70: β = -0.005, SE = 0.022, t = -0.23, p = 0.82) (Male - APOE ε22/23 bar in dashed purple). Detailed statistics for this model can be found in **Supplemental Info 2**. Overall, the positive effect of carrying *APOE* ε44 and ε22/23 was sex-dependent, with females showing a strong advantage between 40-50 in both genotypes.

### *APOE* e44 associated with medial temporal lobe volume loss but increased visual cortex volume

We next investigated differences in brain volume across genotypes in both sexes. Multiple regression analysis with Bonferroni correction for 137 multiple comparisons yielded six grey matter regions that significantly differed between carriers of *APOE* ε33/34 and ε44 (**Figure 5)**. Regions with lower volumes in carriers of the ε44 variant were limited to the medial temporal lobe with medium effect sizes; left amygdala (f = 0.17), right amygdala (f = 0.20), left hippocampus (f = 0.10), right hippocampus (f = 0.095) (yellow). Albeit with a smaller effect size (f = 0.0029, 0.037 respectively), both left and right intracalcarine cortices were significantly increased in ε44 carriers (light blue).

**Figure 5:**
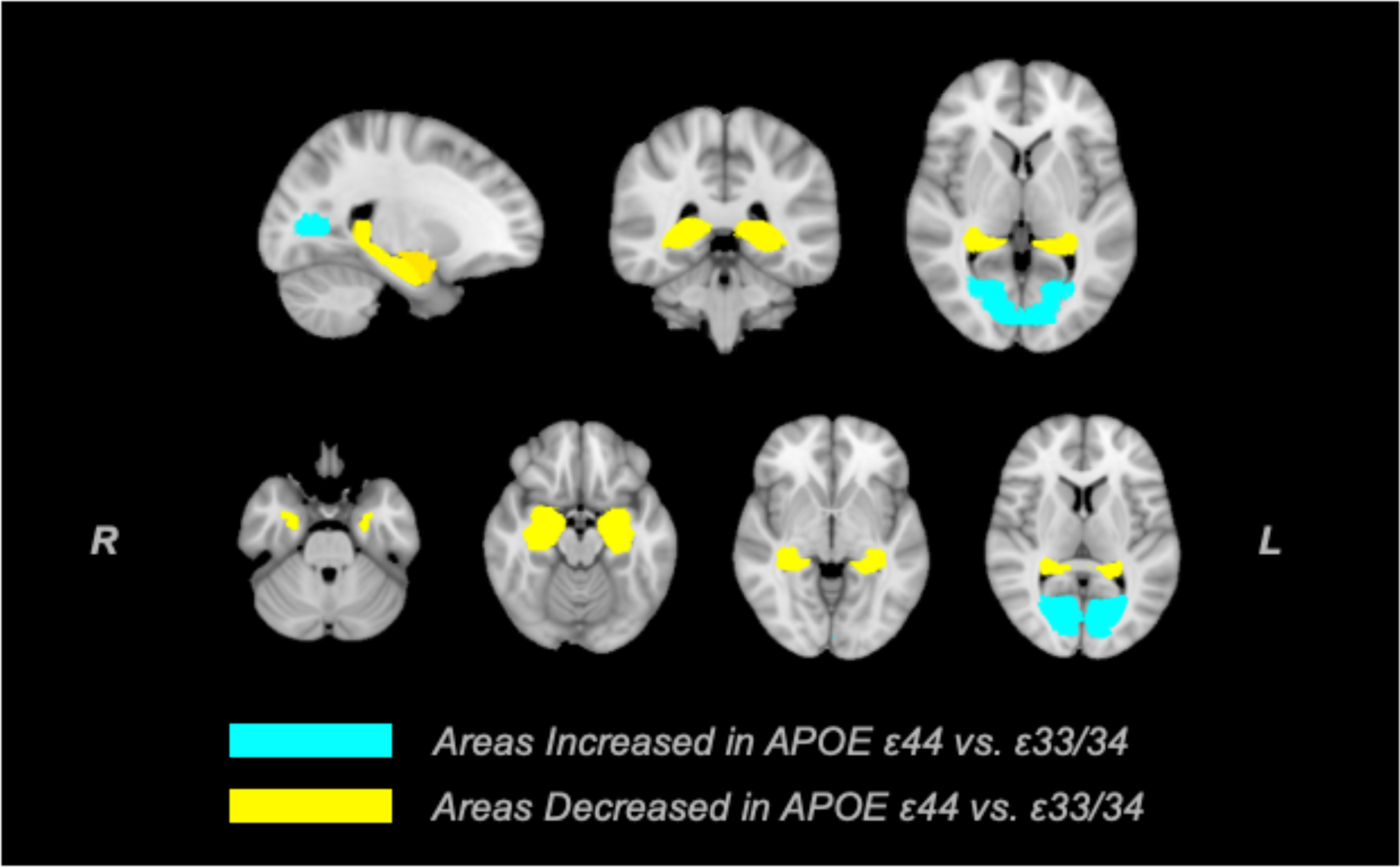
Brain regions modulated by *APOE* ε44 carriers compared to ε33/34 carriers. Brain topography illustrates grey matter regions with significantly increased (light blue) or decreased (yellow) volumes. The top row shows a coronal, sagittal and horizontal section of the brain using the MNI T2 template. The second row shows ventral to dorsal sections of the horizontal section. Region specific decline by age is shown in Figure 6.

**Figure 6:**
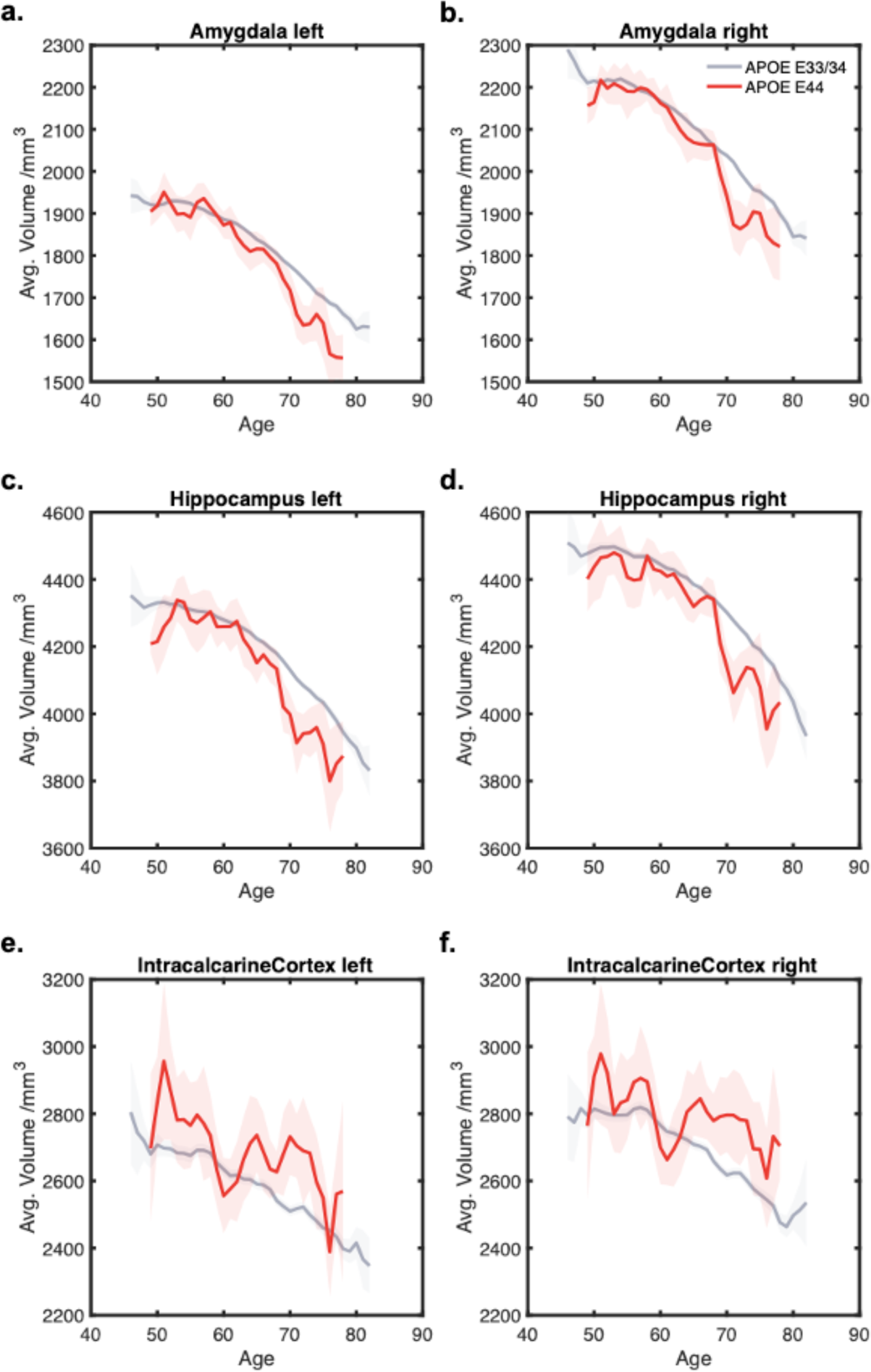
Effect of *APOE* ε44 on Brain Volumes. **Panels a-d:** Significantly reduced brain regions in individuals with the *APOE* ε44 compared to ε33/34. **Panels e-f:** Regions with significantly increased volumes associated with the APOE ε44 compared to ε33/34. These changes in brain volume are visualised using three-year sliding averages. Shaded area represents the SEM for each age group.

A consistent and significant main effect of age was observed across all regions. The main effect of sex was significant for the left amygdala (β = 14.41, SE = 2.41, t = 5.97, p < 0.001) and left hippocampus (β = 3.36, SE = 0.90, t = 3.72, p < 0.001), where these regions were significantly larger in males. No other brain region was significantly modulated by sex. Importantly, there was no female advantage of brain volumes corresponding to the behavioural results. Furthermore, no significant interaction effects between *APOE* genotype and sex were detected across the examined regions. ε22/23 did not have significantly modulated brain regions compared to ε33/34, nor did *Klotho* VS/FC carriers over *Klotho* FC/FC carriers.

### Brain volumes correlate with cognitive scores

Given that six brain regions were significantly different between APOE ε33/34 and ε44, we assessed whether the volumes of these regions correlated with cognitive scores. Both total grey matter volume and the z-score sums of the regions that were significantly different in APOE ε44 showed significant correlations with cognitive scores (**Figure 7**). To determine if the effect of genotype (ε33/34 vs. ε44) on cognition was mediated by brain volumes, a mediation analysis was also conducted. This analysis examined whether the relationship between the APOE ε44 genotype and cognitive decline was mediated by brain volumes that were significantly reduced in APOE ε44 carriers compared to ε33/34 carriers. The results indicated that the ε44 genotype was significantly associated with cognitive decline, and this relationship was partially mediated by brain volumes. Specifically, the mediation analysis revealed that the association between the ε44 genotype and cognitive decline was attenuated when the indirect path involving brain volumes was included. This reduction was evidenced by a modest but significant indirect effect of β = 0.0172 (p = 0.007), as determined by the permutation test (see methods).

**Figure 7:**
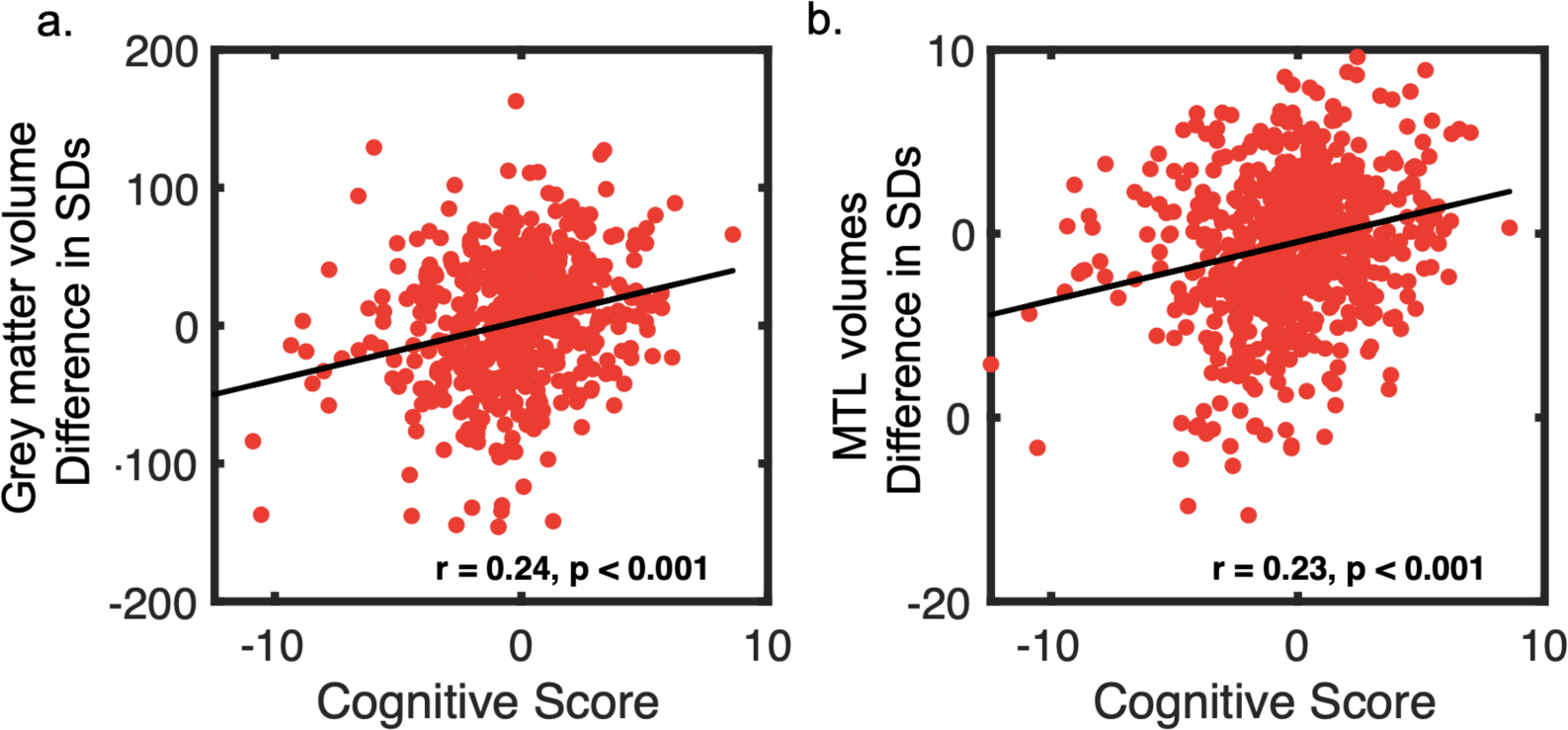
Correlation between brain volume and cognitive score in APOE ε44. **Panels a:** Total grey volumes (standarised units) are z-scored and plotted against an individual’s cognitive score. **Panels b:** Regions that were significantly affected by genotype (bilateral amygdala and hippocampus) plotted against an individual’s cognitive score. Correlations are reported as Pearson’s R.

## Discussion

This study investigated the influence of apolipoprotein (*APOE)* and *Klotho* genotype, alongside the effects of age and sex on cognitive function and brain volume in the largest cohort of healthy individuals examined to date. The analysis revealed differential effects of *APOE* genotypes on cognition, as well as a notable absence of any effect of *Klotho-VS* heterozygosity. The interactions between *APOE* genotype, sex, and age, indicated a complex influence of sex and age on cognition. Most notably, the age-related cognitive trajectories showed an antagonistic pleiotropy effect, whereby the *APOE* ε44 gene associated with a later life disadvantage conferred an advantage at younger ages^23^. However, this effect was limited to younger female carriers of the *APOE* ε44 variant, highlighting the importance of assessing sex-specific preclinical cognitive profiles for the early stages of dementia.

### The deleterious effect of ε44 on cognition emerges with age

*The APOE* ε44 genotype has been highly associated with the pathogenesis of AD^5,6^. Furthermore, A*POE* ε4 homozygosity has been suggested as a new genetic form of AD (Fortea et al., 2024) and its risk in healthy individuals requires further investigation. Our study replicated the well-established decline in cognition of healthy *APOE* ε44 carriers compared to ε33/34 carriers^8–10^. However, we found that in our cohort of healthy participants, ε44 carriers at the study baseline age (40 years) exhibited a cognitive advantage over ε33/34 carriers. This is consistent with APOE ɛ4’s association with enhanced cognitive performance in younger adults^19–22^ and may be attributed to previously reported antagonistic pleiotropy effects^23^. The highest known genetic risk factor for AD is paradoxically associated with earlier advantages with resistance to infection^50^, improved fitness during foetal development^51^, enhanced fertility^50,52^ and here, higher cognitive abilities. These key benefits in early development likely explain the persistence of this AD-risk-conferring gene in our genome.

### Antagonistic pleiotropy of cognition is sex-specific

Cognitive trajectories by age and sex revealed that young female *APOE* ε44 carriers up to 45 years old displayed a cognitive advantage over ε33/34 carriers, whereas males did not. Therefore, this antagonistic pleiotropy effect in cognition was specifically driven by females. Although a general female advantage in their 40s has been previously shown in cognition^13^, the data presented here show that this female advantage is pronounced specifically in *APOE* ε44 carriers. The ε44 female advantage in cognitive performance diminished with age, with males exhibiting higher cognitive abilities from mid-life onward, consistent with previous findings that males exhibit higher cognitive abilities at this age range^30^. This finding is also supported by studies showing that female ε44 carriers are more likely to go on to develop AD^28^, have greater atrophy of the hippocampus^17^ and are at a higher risk of AD pathology^53^. Furthermore, these results are aligned with a meta-analysis assessing the link between APOE ε4 and sex differences, reporting that age differentially affected the risk of developing MCI and AD, with an increased risk of developing AD in women between 65 and 75 years of age^27^. Therefore, age strongly impacts the sex-mediated decline in cognition, where we identify both an early-age advantage and a later-age decline. Further assessment of the mechanisms for age-related effects in females may be relevant to assess in terms of the recently characterized tau pathology in females^54–56^. Given the impact of sex on brain health concerning AD, exploring sex-specific therapies might help to elucidate mechanisms underlying sex-related vulnerabilities across different ages^57^.

### APOE ε44 effects on brain structure

The analyses performed here also revealed that carrying ε44 was associated with smaller hippocampi and amygdala. These grey matter volume differences also correlated with the cognitive score and mediated the effect of ε44 on cognition, suggesting a potential structural basis for the cognitive decline observed in these individuals in later life. Given the time interval between cognitive testing and MRI scans (∼10 years) such small but significant correlations reflect the potential association between brain anatomy and cognition. These findings are aligned with studies showing that genetic predispositions, especially APOE ε44, can affect the hippocampus in healthy individuals^15,58^. In this cohort, ε44 carriers also had enlarged bilateral intracalcarine cortices, consistent with a previous study on the same cohort reporting that the intracalcarine cortex correlated with the number of copies of ε44 alleles^59^. While this dataset does not allow for causal inference, one potential explanation may be a compensatory recruitment process. For example, frontal activation in ε44 carriers has been discussed in relation to AD-related processes^23,24,60^. Such a potentially protective or compensatory mechanism may enlarge the intracalcarine cortices in this genotype. This feature strongly associated with the visual cortex may also relate to the superior cognitive performance observed in ε44 carriers^22,61^, which is reported to last through aging in some visual domains^62^. The analyses conducted here also showed an enlarged left amygdala and left hippocampus males compared to females. These brain areas are important regions related to the neuropathological lesions of AD^63^. No significant association of female early advantage could be identified in grey matter volumes, although previous studies have shown reduced functional connectivity to the hippocampus in female *APOE* ε4 carriers^16^. Finally, no antagonistic pleiotropy effects were found in brain volumes, likely owing to the limited sample size that prevented equivalent analysis of age-related decline.

### APOE ε22/23 effects

The effects of the *APOE* ε2 variant were also assessed in comparison to the reference group of *APOE* ε33/34. The ε2 allele is associated with a lower risk of AD^64^ and increased longevity^65^. ɛ2 carriers are also more likely to live up to 100^66^, but the cognitive trajectory across aging has not been clearly defined. When comparing the effects of carriers of the ε22/23 genes, we found that ε2 carrier advantages became evident with aging. The cognitive decline with age was less steep compared to the reference ε33/34 carrier group. This highlights the potential protective effect of this allele, especially with age. Sex also modulated the effects whereby the positive effects of ε22/23 were higher in females at younger ages but changed to a male advantage with aging as was the case for ε44 carriers. Although some previous studies have reported increased cortical thickness^67^ and larger hippocampal volume in healthy elderly ɛ2 carriers^68^, reports are mixed with others showing no differences^69^. Here, we also showed no difference in hippocampal volume.

### Absence of Klotho effect

While extensive research in both mice^33,34^ and humans^36,38^ has explored the benefits of *Klotho*- VS heterozygosity, this study did not reveal any significant effects of *Klotho*-heterozygosity. Previous research suggested that individuals with the *Klotho*-VS/FC genotype displayed elevated neuropsychological test scores, as indicated by a global composite score^38^. However, here the potential cognitive advantages associated with *Klotho* heterozygosity were not observed. Moreover, no effect on brain grey matter volumes was observed, which contrasts with previous findings indicating improved memory and executive functions in *Klotho* heterozygotes, correlated with a larger prefrontal cortex^40^. Furthermore, *APOE* status did not modulate *Klotho* effects and therefore did not show the hypothesised, neuroprotective effects of *Klotho* VS/FC, contrary to prior findings^44,45^. However, the results of at least one investigation concluded that *Klotho* VS/FC does not modify amyloid beta burden in individuals with *APOE* ε44^47^, which is aligned with the findings from this dataset.

One potential explanation for mixed findings in the literature is the lack of a direct correlation between *Klotho* genotype and Klotho protein levels. Recent studies demonstrating the beneficial effects of Klotho secretion have directly evaluated Klotho levels through injection^70^ or quantification^41^. Secreted serum Klotho may benefit brain health, but in population-based cohort studies that solely investigate genotype, we may lack the sensitivity of quantifying protein levels, which may be highly influenced by transcription processes. Klotho secretion is also highly modulated by environmental factors like chronic stress^71^, which unlike socioeconomic status, is uncorrected for in this study. Further characterisation of the Klotho protein levels will be important to understand the genetic effects of *Klotho* on brain and behaviour and assess its therapeutic benefits on neurodegeneration and longevity.

### Importance of large sample sizes

Understanding the progression of neurodegeneration requires assessing early changes preceding clinical diagnosis, a task facilitated by the UK Biobank data. The impact of genes has often been overshadowed by a focus on disease associations rather than risk factors in the healthy population. Here, we offer a new perspective by investigating genetic risk factors, age and sex in a large number of healthy participants. This cohort is the largest sample of *APOE* ε44 carriers to date, which allows us to investigate varying factors influencing brain and behaviour. Furthermore, the characterisation of trajectories over time is a critical aspect of the analysis, which has been overshadowed in previous investigations assessing the interactions of Klotho and APOE genes of aging^46^. Inconsistencies in previous research regarding cognitive advantages among ε4 carriers may stem from limited statistical power, obscuring assessments of sex and age-related effects.

### Study limitations

Although the UK Biobank provides a valuable resource for studying cognitive impairment, several limitations warrant discussion. Firstly, participants tend to be healthier and of a higher socioeconomic status than the general population^72^, potentially underrepresenting those at higher risk for cognitive impairment. Secondly, the 10-year average difference between behavioural testing and imaging data collection challenges the correlation of brain and behaviour. Thirdly, our analysis reduced various cognitive tasks into a composite score using principal component analysis, and therefore overlooked the variations across different cognitive domains. This analysis method was motivated by the lack of validated domain sensitivity of the tasks administered in the UK Biobank. While general cognition is important to study, specific cognitive domains should also be considered in future studies with pre-defined hypotheses, as some domains of cognition may be differentially affected. For example, in the context of *APOE*, short-term and long-term retention has been shown to exhibit distinct effects and may warrant separate consideration^61^. Improved cognitive testing which is sensitive to subtle cognitive deficits may facilitate this line of research. Finally, as this cohort is made of healthy individuals, clinical conversion is not assessed, which may be critical to assess as not all cognitive decline in healthy is associated to pathological states. Notwithstanding these limitations, our study’s broad age range and large sample size provide a strong foundation for future research on how to better stratify those at risk of developing dementia.

### Conclusions

This study leveraged one of the most extensive genotyped cohorts of healthy individuals from the UK Biobank. The complex interplay between *APOE*, sex, and age, contributes to our understanding of how genetic effects are highly dependent on sex and age in both carriers of *APOE* ε4 and ε2. The female-specific antagonistic pleiotropy effects are a novel finding that highlights the importance of assessing sex-dependent cognitive trajectories of dementia. Finally, despite the absence of significant effects from *Klotho*-VS heterozygosity on cognition, the mechanisms of *Klotho* on cognition still warrant investigation. Protective genetic factors that may modify the deleterious effects of *APOE* ε4 in later life remain a critical target for the development of therapies and diagnostics against aging and neurodegeneration.

## Methods

### Participants

The participants in this study were drawn from the UK Biobank (https://ukbiobank.ac.uk), a population-based prospective cohort of healthy adults. 320,861 participants between the ages of 40 and 70 years were included in this study. Individuals with a history or current diagnoses of the following were excluded from the study: AD or Parkinson’s disease, stroke, brain/head injury, transient ischaemic attack, subdural or subarachnoid haematoma; infection of the nervous system; brain abscess, haemorrhage, skull fracture, encephalitis, amyotrophic lateral sclerosis, meningitis, chronic neurological problem, multiple sclerosis and epilepsy. Those with alcohol, opioid and other dependencies were also removed from the analysis. Follow-up self-reported questionnaires and hospital records were used to assess inclusion in the study. Participants were genotyped during the data collection phase. Rarer variants of *APOE* (ε1) and *Klotho* (homozygous (VS/VS) were not included in the analysis. The effects of carrying the APOE ε44 genotype were studied by comparing them to ε33 and ε34 carriers. For further exploratory analysis, we also compared ε2 carriers (ε22 and ε23) to the same reference (ε33 and ε34 carriers). The homozygous ε4/ε4 genotype was analysed relative to the reference as this group is recognised for having the most significant impact on cognition and brain changes, and a potential genetic form of AD^7^. The scarcity of homozygous carriers has limited this grouping; however, this limitation was overcome by the large sample size provided by the UK Biobank. For *Klotho*, the *Klotho*-VS heterozygous variant (FC/VS) was compared to the most common *Klotho* variant (FC/FC). 37,976 individuals had brain MRI data which was used to analyse differences in grey matter volume. Data was acquired from an initial baseline visit between 2006 and 2010, and the imaging visit occurred between 2014 and 2020. All participants provided written, informed consent and the study was approved by the Research Ethics Committee (REC number 11/NW/0382).

### Cognitive testing

Cognitive testing was performed via fully automated touchscreen tasks. We analysed nine short cognitive tasks covering a range of cognitive domains, including processing speed, episodic memory, working memory and reasoning (**Table 1**). High validity and test-retest reliability of these UK Biobank tasks have been reported previously^73^.

**Table 1:**
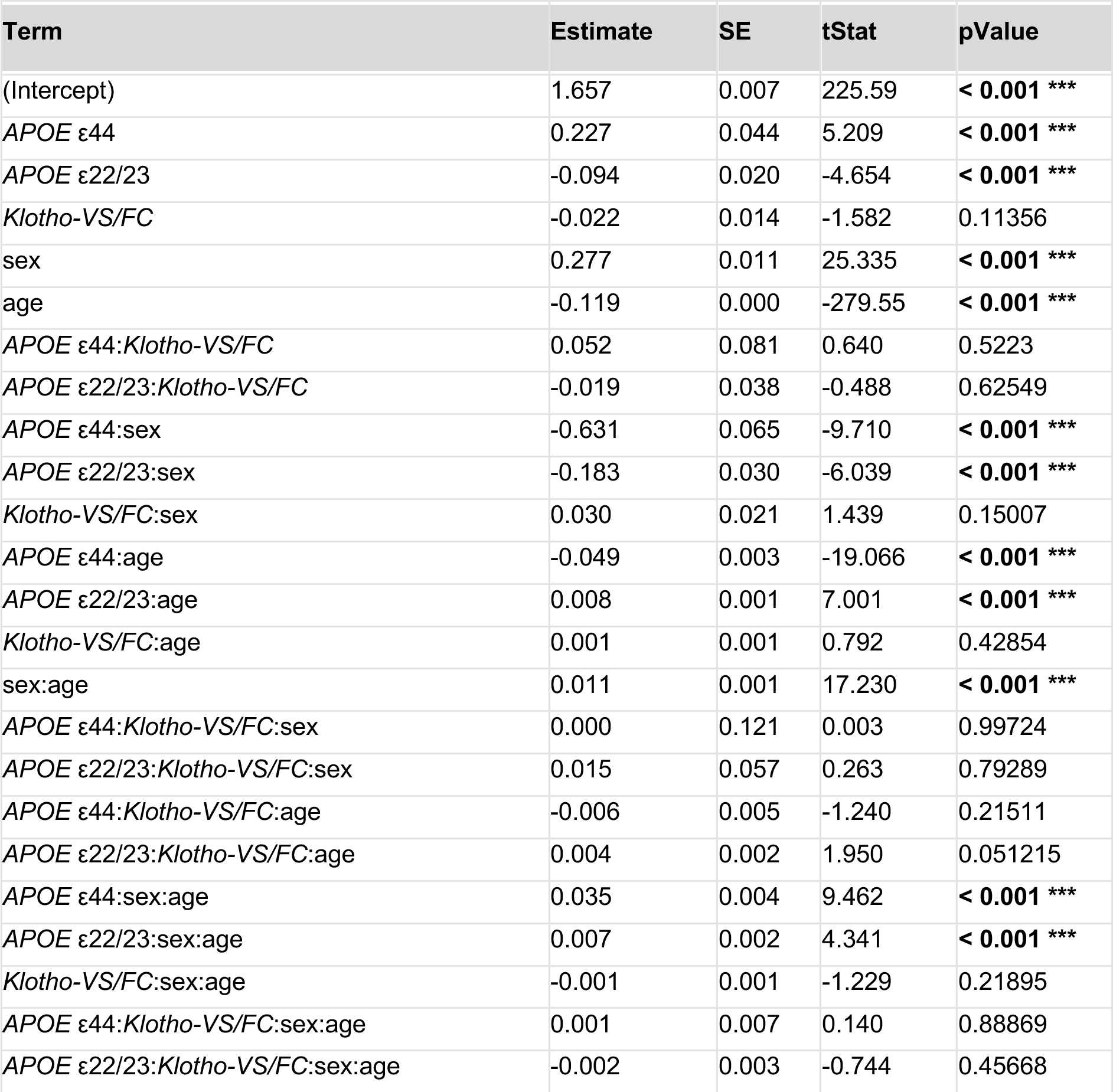
Regression model results for the effect of *APOE, Klotho*, sex and age. Results of the multiple regression analysis, incorporating predictor: APOE (ε33/34, ε44, ε22/23), Klotho (FC/FC, FC/VS), sex, age, and all their possible interactions. Each row represents a predictor or interaction, with significant p-values highlighted in bold. The corresponding coefficients and standard errors (SE) are visually represented in Figure 3, Panels a & b.

| Term | Estimate | SE | tStat | pValue |
| --- | --- | --- | --- | --- |
| (Intercept) | 1.657 | 0.007 | 225.59 | **< 0.001 \*\*\*** |
| *APOE* ε44 | 0.227 | 0.044 | 5.209 | **< 0.001 \*\*\*** |
| *APOE* ε22/23 | -0.094 | 0.020 | -4.654 | **< 0.001 \*\*\*** |
| *Klotho*-VS/FC | -0.022 | 0.014 | -1.582 | 0.11356 |
| sex | 0.277 | 0.011 | 25.335 | **< 0.001 \*\*\*** |
| age | -0.119 | 0.000 | -279.55 | **< 0.001 \*\*\*** |
| *APOE* ε44:*Klotho*-VS/FC | 0.052 | 0.081 | 0.640 | 0.5223 |
| *APOE* ε22/23:*Klotho*-VS/FC | -0.019 | 0.038 | -0.488 | 0.62549 |
| *APOE* ε44:sex | -0.631 | 0.065 | -9.710 | **< 0.001 \*\*\*** |
| *APOE* ε22/23:sex | -0.183 | 0.030 | -6.039 | **< 0.001 \*\*\*** |
| *Klotho*-VS/FC:sex | 0.030 | 0.021 | 1.439 | 0.15007 |
| *APOE* ε44:age | -0.049 | 0.003 | -19.066 | **< 0.001 \*\*\*** |
| *APOE* ε22/23:age | 0.008 | 0.001 | 7.001 | **< 0.001 \*\*\*** |
| *Klotho*-VS/FC:age | 0.001 | 0.001 | 0.792 | 0.42854 |
| sex:age | 0.011 | 0.001 | 17.230 | **< 0.001 \*\*\*** |
| *APOE* ε44:*Klotho*-VS/FC:sex | 0.000 | 0.121 | 0.003 | 0.99724 |
| *APOE* ε22/23:*Klotho*-VS/FC:sex | 0.015 | 0.057 | 0.263 | 0.79289 |
| *APOE* ε44:*Klotho*-VS/FC:age | -0.006 | 0.005 | -1.240 | 0.21511 |
| *APOE* ε22/23:*Klotho*-VS/FC:age | 0.004 | 0.002 | 1.950 | 0.051215 |
| *APOE* ε44:sex:age | 0.035 | 0.004 | 9.462 | **< 0.001 \*\*\*** |
| *APOE* ε22/23:sex:age | 0.007 | 0.002 | 4.341 | **< 0.001 \*\*\*** |
| *Klotho*-VS/FC:sex:age | -0.001 | 0.001 | -1.229 | 0.21895 |
| *APOE* ε44:*Klotho*-VS/FC:sex:age | 0.001 | 0.007 | 0.140 | 0.88869 |
| *APOE* ε22/23:*Klotho*-VS/FC:sex:age | -0.002 | 0.003 | -0.744 | 0.45668 |

**Table 2:** Cognitive tasks of the UK Biobank. Nine cognitive tasks from the UK Biobank were used to construct a composite cognitive score.

#### Reaction Time Task

This task measured reaction time to visual stimuli using a modified Go/No-Go format. The mean time taken to respond to matching pairs was used as the measure of reaction time.

#### Trail-Making Task

Trail-making assessed executive function. Trail A required participants to click on numbers in ascending order. Trail B required participants to click on numbers and letters in an alternating and ascending pattern (e.g., 1-A-2-B-3-C). The difference in completion time between Trail A and B was used as an index of executive function.

#### Pairs Matching Task

This task assessed visual memory. Participants memorised the position of as many matching pairs of cards arranged randomly on a grid. The cards were then turned face down and participants were asked to select as many pairs as possible in the fewest tries. The number of incorrect matches until all matches were made was used as the outcome measure.

#### Fluid Intelligence Task

This task measured IQ by probing verbal and numerical reasoning. Participants answered 12 multiple-choice questions (between 3 and 5 choices) that examined logic and reasoning ability. We used the fluid intelligence score to index their performance.

#### Paired-associative Task

This task measured verbal declarative memory. Participants memorised word pairs displayed for 30 seconds. After a delay (Matrix Pattern Completion Task) one word of the pair was presented and the corresponding pair had to be selected from a choice of four options. The outcome variable was the number of correctly answered pairs.

#### Tower Task

This task tested planning abilities. Participants were shown 2 displays of 3 pegs and 3 different coloured hoops placed on the pegs. Participants counted how many moves it would take to go from one configuration of hoops to the other. The outcome variable was the number of items answered correctly in 3 minutes.

#### Symbol-digit Substitution Task

This task measured processing speed. Participants were presented with a key that paired symbols with numbers. Participants were shown a row of symbols and entered the associated number using a keypad. The score was the number of correct symbol-digit matches made in 60 seconds.

#### Matrix Pattern Completion Task

This task tested non-verbal fluid reasoning. Participants were presented with a matrix design, with a missing piece in the lower right corner. Participants identified what the missing piece would look like from a list of options. The outcome variable was the number of items answered correctly in 3 minutes.

#### Numeric Memory Task

Short-term memory and working memory capacity was tested using a backward digit span task. Participants were simultaneously presented with a sequence of numbers and were asked to recall and repeat the numbers in reverse order. The outcome variable was the maximum number of digits correctly remembered in the reverse order.

### Behavioural analysis

Analyses were carried out on MATLAB R2021b. To examine the effect of genotype on cognitive functions, behavioural measures of each task were first z-scored. For tasks where lower values denoted superior performance (i.e. reaction time task, trail-making task, and pair-matching task), the values were adjusted so that negative z scores reflect below-average performance. Missing values were extrapolated using a matrix based on the age, sex, and *APOE* genotype of this participant. Principal component analysis (PCA) was applied to these scores to reduce dimensions and establish a composite score of cognition. The 1st principal component was taken as the composite score of cognition for each individual. These scores were corrected for socioeconomic status using the Townsend deprivation index and years of education.

We investigated the factors affecting cognition using multiple linear regression, modelling cognition as a function of *APOE*, *Klotho*, sex and age, and their interactions.

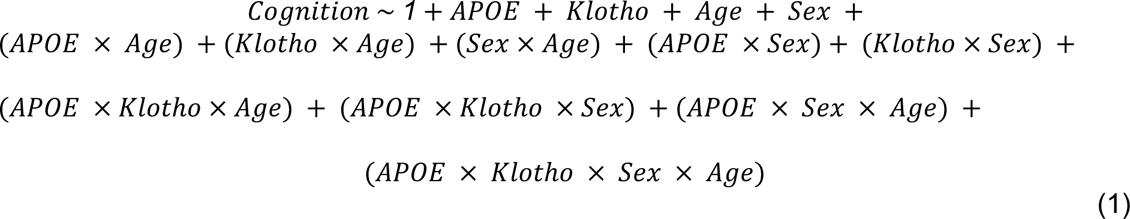

For the purpose of this analysis, the reference genotype was set to the most common grouping of variants of each of the genes studied; *APOE* ε33/34 and *Klotho* FC/FC. The reference sex was set as female. The minimum age was subtracted from all ages so that all main effects and interaction terms without the age predictor represent an effect at the minimum age in the data of 40 (*APOE* ε33/34: n = 3361, *APOE* ε44: n = 129, *APOE* ε22/23: n = 513). This regression model explained 44.2% of the variance in cognitive scores (R-squared = 0.442), with a highly significant fit (F = 12,700, p < .001). We used the Akaike Information Criterion (AIC) and Bayesian Information Criterion (BIC) to confirm the superiority of this complex model over a model without interactions between predictors. The full model had a lower AIC (AIC: 945,900) compared to the simple multiple regression model (AIC: 947,490) with no interactions and explained 42.2% of the variance compared to 41.9% variance. The model incorporating all interactions revealed a significant age effect, prompting further analysis of the directionality of *APOE* and sex across three age brackets: 40-50, 50-60, and 60-70 years. This segmentation equally divides the age ranges into three groups to assess whether the directionality of main effects changes depending on age. As *Klotho* did not influence the results, its effect was excluded from these subsequent age-binned regressions. Each age group was analysed using the below regression model:

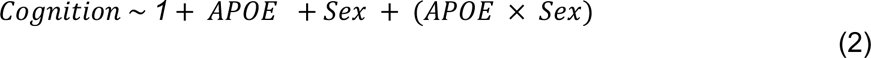

### Imaging

Magnetic resonance imaging (MRI) data were acquired on Siemens Skyra 3T scanner as part of the UK Biobank neuroimaging protocol, on average around 10 years after the initial assessment. 1 mm isotropic voxel high-resolution T1-weighted images were collected and pre- processed using a standard previously published pipeline implementing quality control^74^. For volumetric analysis of T1-imaged structures, gradient distortion correction and registration onto MNI152 space were applied. FAST (FMRIB’s Automated Segmentation Tool) was used to segment white matter, grey matter, and cerebrospinal fluid. We extracted 137 imaging-derived phenotypes (IDPs) as key grey matter structures. Volumes were corrected for years of education, socioeconomic status, head size, scan date and table position using a general linear model. We examined the difference in grey matter volumes using a whole-brain approach across genotypes. We ran a multiple linear regression for each brain region and reported the brain regions that had a significant main effect of genotype. To correct for the multiple comparisons made across all brain regions, we applied Bonferroni correction with an adjusted alpha threshold of 0.05/137. Given the substantially smaller sample size and multiple regressions needed to be run for whole brain analysis, linear models which included interactions did not show any significant main effect or interaction effects. We therefore assessed the additive effects of genotype, sex and age independently on grey matter volume using the following regressions. Regression models for ε44 and ε22/23 were assessed separately. Cohen’s f was used as a measure of effect size.

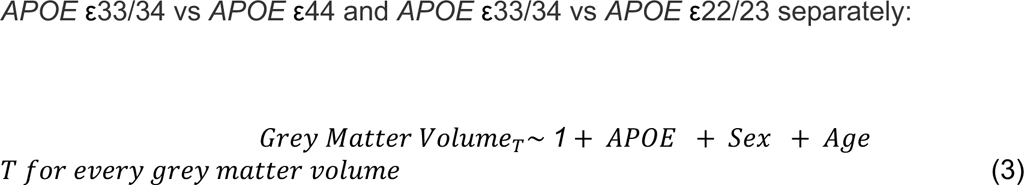

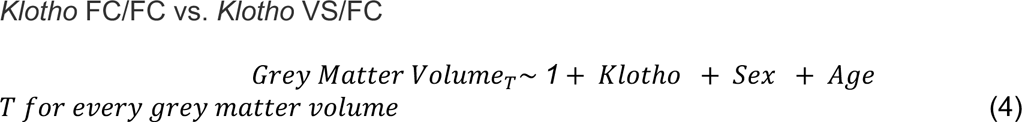

### Brain behaviour correlations

All brain volumes used for brain behaviour correlation analysis were z-scored. A mediation analysis was conducted to examine whether the reduction in brain volumes mediates the relationship between the APOE ε44 genotype and cognitive decline. For the purpose of this hypothesis driven analysis, participants with the APOE ε22/23 genotype were excluded, resulting in a binary APOE variable (ε44 vs. ε34/33). Volumes that were significantly reduced in carriers of APOE ε44 were z-scored and summed to create a composite measure of medial temporal lobe volume reduction. Logistic regression (APOE ∼ Volume) and linear regressions (Cognitive Score ∼ Volume and Cognitive Score ∼ Volume + APOE) were performed to estimate the coefficients for the mediation model. The observed indirect effect (a * b) was calculated as the product of the coefficients from these models. A permutation test with 1000 permutations was conducted to assess the significance of the indirect effect by permuting the APOE variable and recalculating the indirect effect for each permutation. The p-value was determined by comparing the observed indirect effect to the distribution of permuted indirect effects.

## Data availability

UK Biobank data is openly available at www.ukbiobank.ac.uk

## Acknowledgements

This work was funded by Wellcome Trust Principal Research Fellowship to MH, the MRC Clinician Scientist Fellowship [MR/P00878X] and NIH Oxford Biomedical Research Centre to SGM, the Wellcome Trust PhD clinical fellowship to XT and the Berrow Foundation Scholarship to KS. The authors would like to thank Dr. Sofia Toniolo and Dr Sijia Zhao for their comments on the manuscript. For the purpose of open access, the author has applied a CC BY public copyright licence to any Author Accepted Manuscript version arising from this submission.

